# Inter-rater reliability of the instrumented lean-and-release (iLEAN) test for manual assessment of reactive balance: A pilot study in simulated older adults

**DOI:** 10.1101/2025.09.10.25335475

**Authors:** Tatsuya Hirase, Asahi Nishimura, Yuika Kato, Nanako Ishizaka, Naoya Hasegawa, Stephen R. Lord, Yoshiro Okubo

## Abstract

**Background and Purpose:** Quantitative assessment of reactive balance often requires large, costly equipment, limiting its use in clinical practice. This study investigated the inter-rater reliability of the instrumented lean-and-release (iLEAN) test, a manual reactive balance assessment designed to provide a practical alternative for physiotherapists.

**Methods:** Sixteen healthy young women wore knee braces, weighted vests, and ankle weights to simulate reduced physical function typical of older adults. Participants completed the iLEAN test in standing, where reactive balance control was assessed in forward, left, right, and backward directions. During tests, an assessor applied the iLEAN device against the shoulder and released support once the participant reached a target lean load, which was progressively increased until balance could not be regained with a single step. Inter-rater reliability was analyzed using intraclass correlation coefficients (ICC, model 2,1) and Bland–Altman plots.

**Results:** The ICCs (95% confidence intervals) for the iLEAN scores were 0.60 (0.17– 0.84) for forward, 0.75 (0.34–0.92) for left, 0.80 (0.45–0.94) for right, and 0.83 (0.58– 0.94) for backward. Bland–Altman analysis revealed no systematic bias between raters across all directions.

**Discussion:** The iLEAN demonstrated acceptable inter-rater reliability in all directions, with the backward test showing the highest consistency. These findings suggest the iLEAN, particularly in the backward direction, may provide physiotherapists with a reliable and practical method to monitor changes in reactive balance control during exercise or rehabilitation, without reliance on specialized laboratory equipment.

## Introduction

Falls in community-dwelling older adults are often precipitated by postural perturbations, such as trips and slips during walking (Berg et al., 1997). Reactive balance—the ability to restore posture following a sudden mechanical disturbance—is a critical determinant of fall prevention (Mohamed Suhaimy et al., 2021). Evidence from systematic reviews indicates that older adults with a history of falls exhibit impaired reactive stepping responses compared to their non-falling peers (Okubo et al., 2021).

In clinical practice, reactive balance is typically assessed with the pull test or the push-and-release test. These methods are low-cost and have demonstrated good reliability in populations such as people with Parkinson’s disease and polymyositis (Jacobs et al., 2006). However, their precision is limited due to variability in perturbation delivery and the qualitative nature of response scoring. Conversely, the lean-and-release test offers a more standardised and quantifiable assessment of reactive stepping (Inness et al., 2015), but its implementation in routine practice is hindered by the need for specialised equipment, large space, and technical expertise.

To address this gap, we developed the instrumented lean-and-release (iLEAN) test, which incorporates two hand-held dynamometers to provide a portable, manual, and quantitative assessment of reactive stepping in forward, backward, and lateral directions. The purpose of this preliminary study was to evaluate the inter-rater reliability of the iLEAN test in healthy young adults under simulated age-related physical function constraints.

## Methods

### Participants

Sixteen healthy women were recruited through university bulletin board advertisements. Inclusion criteria were absence of neurological, musculoskeletal, or psychiatric disorders. Participants had a mean age of 20.7 years (SD 0.8). The study was approved by the Office of Research Ethics at Kanagawa University of Human Services (Approval No. 18-23-34) and complied with the Declaration of Helsinki. All participants provided written informed consent.

### Demographics

Baseline measures included age, sex, height, weight, body mass index, and dominant leg (Table 1). Dominant leg was identified by asking: “Which leg would you use to kick a ball?” (van Melick et al., 2017)

**Table 1.** Participant characteristics

| Items |  |
| --- | --- |
| Age (years) | 20.7 ± 0.8 |
| Height (cm) | 158.5 ± 5.3 |
| Weight (kg) | 52.4 ± 7.4 |
| BMI (kg/m<sup>2</sup>) | 20.8 ± 2.0 |
| Dominant leg, n (%) | 16 (100) |

Data are presented as mean ± standard deviation or number (percentage). BMI: Body mass index

### Reactive balance assessment

Reactive balance was assessed in forward, backward, left, and right directions using the instrumented lean-and-release (iLEAN) test. The device comprised two hand-held dynamometers (μTas, Anima Corp., Tokyo) held by the assessor and a wrist-worn display connected to an Android device running the iLEAN application (Figure 1).

**Figure 1.**
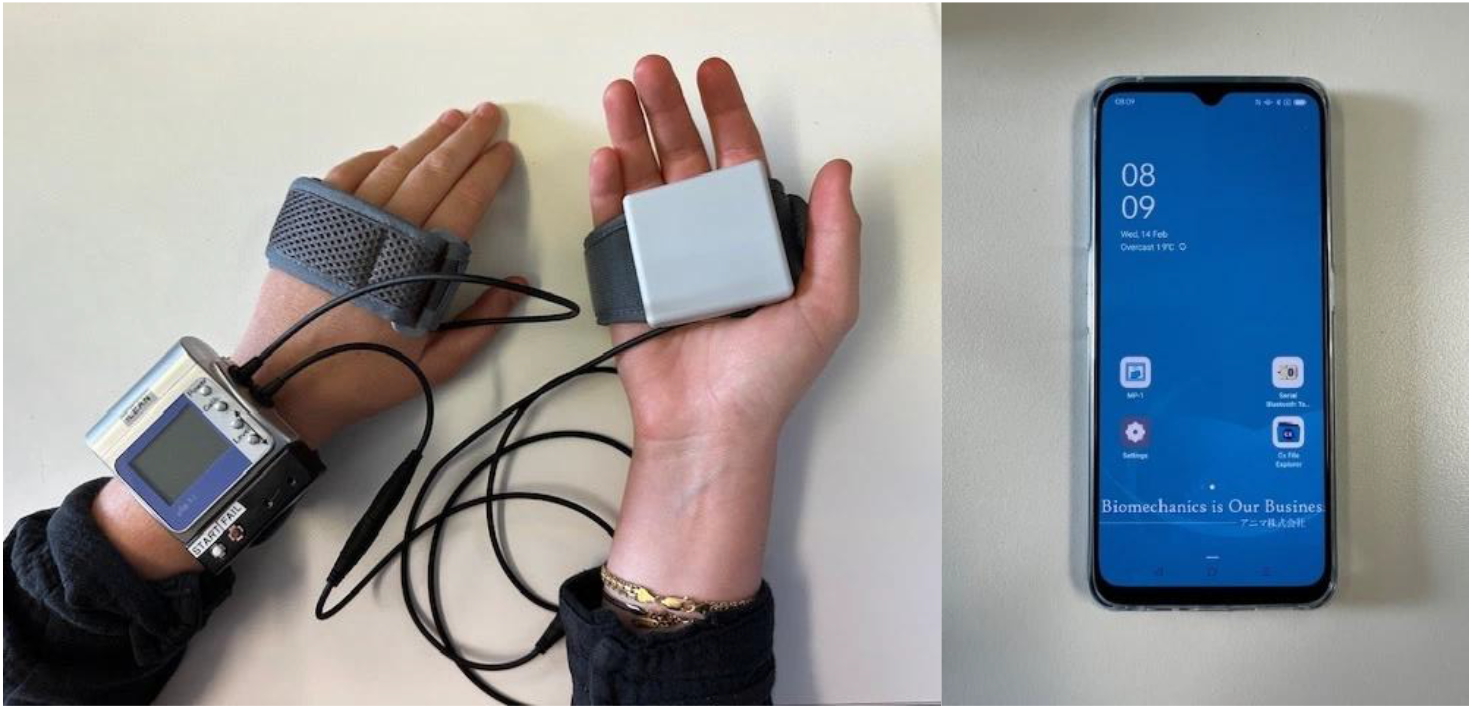
iLEAN device and application

To simulate age-related functional decline, participants wore a 2.7 -kg weighted vest, 1.5-kg ankle weights, and bilateral knee braces (Oitarou, Kyoto Kagaku, Kyoto). For forward and backward trials, participants stood with feet hip-width apart; for lateral trials, feet were together. A side wall was positioned 50 cm away for safety. In forward, left, and right conditions, a foam block (14 cm height, placed 5 cm in front of the stepping foot) was used to prevent recovery with excessively long steps.

For each trial, participants folded their arms across the chest, leaned against the device until a target load was reached, and were then released by the assessor within 1 –5 seconds. The initial load was set at 5% of body weight and increased in 1% increments if balance was recovered with a single step in two consecutive trials. The maximum safe load was capped at 20% of body weight. A trial was considered unsuccessful if participants: (1) unfolded arms, (2) touched the wall, (3) took more than one step, (4) touched the floor with a knee, or (5) in forward/lateral directions, displaced the foam block. The highest load successfully recovered was recorded as the iLEAN score (% body weight).

Two trained physical therapy students (A.N. and Y.K.) served as assessors. Order of assessors was randomized, and assessors were blinded to each other’s results.

### Statistical analysis

Inter-rater reliability for iLEAN scores in each direction was evaluated using intraclass correlation coefficients (ICC, model 2,1) with 95% confidence intervals (CI). ICCs were interpreted as: <0.50 = poor, 0.50–0.75 = moderate, 0.75–0.90 = good, and >0.90 = excellent (Koo & Li, 2016). Bland–Altman plots were used to assess systematic bias. Fixed bias was identified when the 95% CI of mean differences did not include zero, and proportional bias was determined from significant correlations between mean scores and differences. Minimal detectable change at 95% confidence (MDC95) was calculated as: SEM × √2 × 1.96 (Muir-Hunter et al., 2015).

Sample size requirements were estimated using Walter, Eliasziw, and Donner’s (Walter et al., 1998) method, indicating that 12 participants would be sufficient to detect an ICC of 0.9 with a lower CI of 0.60 (α = 0.05, β = 0.20). Statistical analyses were conducted using SPSS 26.0 for Windows (IBM Corp., Armonk, NY, USA).

## Results

Table 2 presents the iLEAN scores and ICCs for each direction. Scores could not be obtained for four participants in the left and right conditions because they were unable to complete the trials at the initial target load without striking the foam block. The ICC (2,1) values were 0.60 (95% CI: 0.17–0.84) for forward, 0.75 (95% CI: 0.34– 0.92) for left, 0.80 (95% CI: 0.45–0.94) for right, and 0.83 (95% CI: 0.58–0.94) for backward directions.

**Table 2.** iLEAN scores and inter-rater reliability

| Directions | iLEAN scores (%) |  | ICC (2,1) | 95%CI of ICC (2,1) |
| --- | --- | --- | --- | --- |
|  | Rater A | Rater B |  |  |
| Forward | 12.4 ± 3.0 | 12.6 ± 3.4 | 0.60 | 0.17 to 0.84 |
| Left | 7.5 ± 2.5 | 7.8 ± 2.1 | 0.75 | 0.34 to 0.92 |
| Right | 8.4 ± 2.2 | 8.7 ± 2.4 | 0.80 | 0.45 to 0.94 |
| Backward | 9.3 ± 2.1 | 9.8 ± 1.9 | 0.83 | 0.58 to 0.94 |

Data are presented as mean ± standard deviation.
ICC; Intraclass Correlation Coefficients, CI; Confidence Interval.

Bland–Altman plots (Table 3, Figure 2) showed that most data points lay within the limits of agreement (±1.96 SD of the differences between raters), with no evidence of fixed or proportional bias across directions. The MDC _95_ values were 2.07 (forward), 1.42 (left), 1.37 (right), and 0.85 (backward).

**Table 3.**
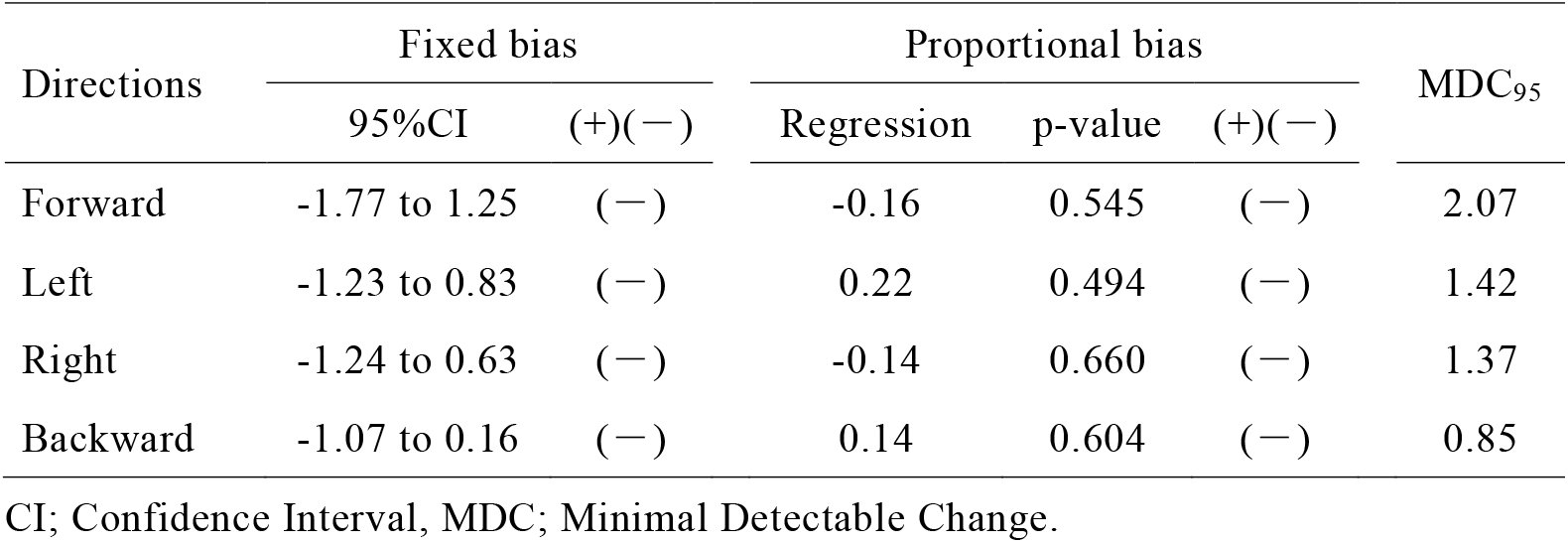
Bland-Altman analysis

**Figure 2.**
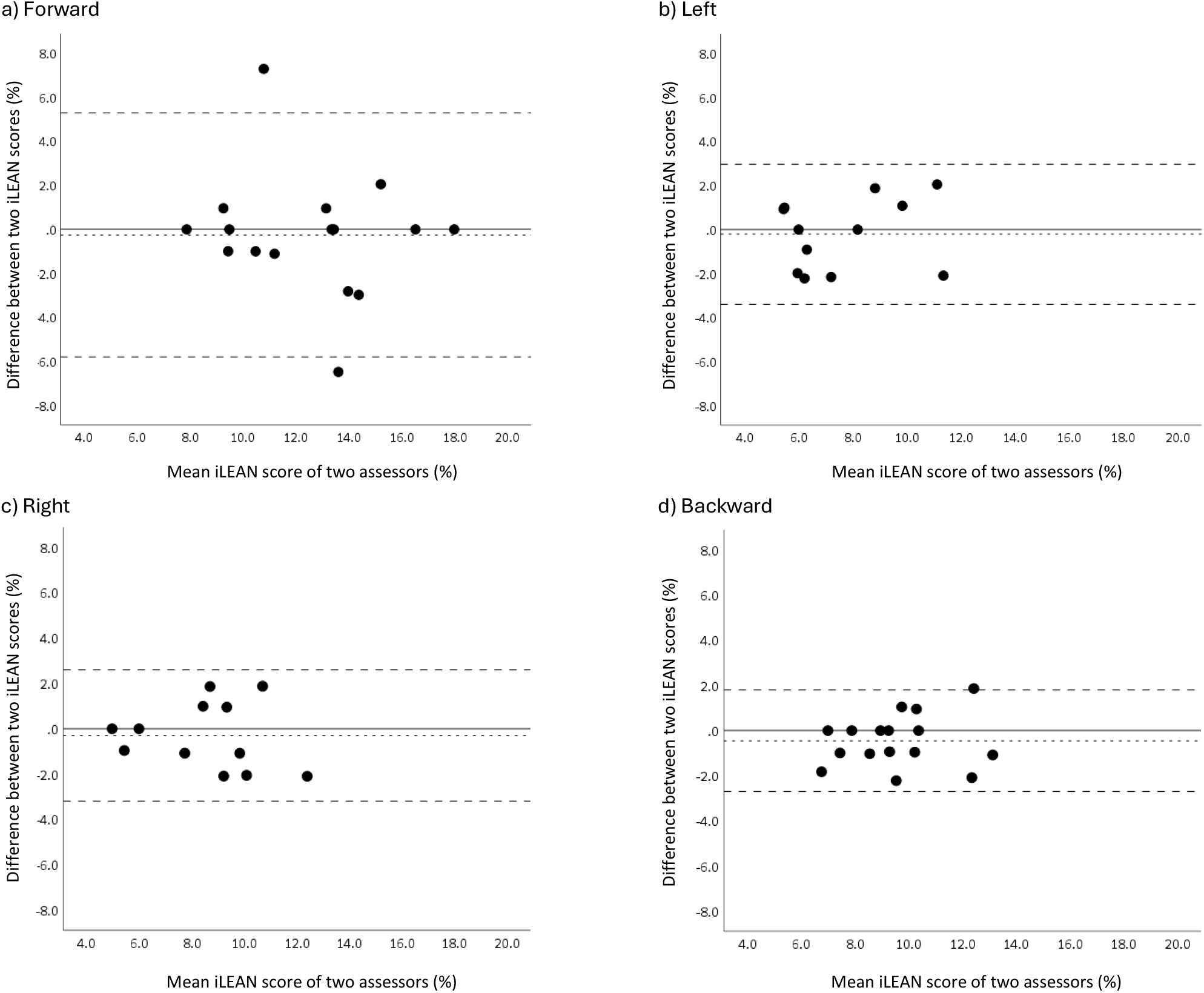
Bland-Altman plot of iLEAN tests Vertical axis is the difference of each value between two assessors for each participant and horizontal axis is the average of each measurement of two assessors. Dotted line shows the mean of differences of the values of two assessors ± 1.96×SD of the differences of the values of two assessors.

## Discussion

This pilot study evaluated the inter-rater reliability of the iLEAN test for assessing reactive stepping ability in healthy young adults. All directions demonstrated acceptable reliability, with the backward condition showing the highest ICC and the smallest MDC_95_ (0.85). Clinically, this suggests that even small changes in backward iLEAN scores may represent meaningful improvements in reactive balance beyond measurement error.

The backward iLEAN test compared favorably with existing manual assessments of reactive balance. Its ICC (0.83) was higher than the pull test (ICC: 0.45 –0.74) (Jacobs et al., 2006) and comparable to the push-and-release test (ICC: 0.83–0.84) (Jacobs et al., 2006). This may reflect the more controlled force application of iLEAN and push-and-release, in contrast to the variable abruptness of the pull test. Importantly, unlike the push-and-release test, which uses ordinal ratings (a 5-point scale from 0 to 4), the iLEAN provides a quantitative measure of reactive stepping capacity, potentially reducing floor and ceiling effects and allowing more sensitive monitoring of change.

In terms of the iLEAN tests in the other directions, the inter-rater reliability in the forward direction was lower (ICC: 0.60, 95% CI: 0.17 to 0.84) than those in other directions. This may be due to the greater leaning loads that assessors were required to support in the forward direction (12.4 ± 3.0%) than other directions (e.g. left: 7.5 ± 2.5%) which may contribute to fatigue and variable holding performance of the assessor during the test. Although the ICCs for the iLEAN tests in the left and right directions showed good inter-rater reliability (ICC: 0.75 for the left direction and 0.80 for the right direction), four participants were unable to complete the initial 5% body weight trials in both the left and right directions due to their stepping foot contacting the foam obstacle. This suggests the obstacle may not be necessary, particularly in clinical populations where balance recovery is likely to be impaired.

Several limitations should be acknowledged. First, the findings from young healthy women may not generalize to older adults or clinical populations. Second, the initial target load of 5% body weight may be too high for frailer individuals, and lower starting loads (e.g., 3%) should be considered. Third, test–retest reliability was not examined. Fourth, only women participated, and further studies should include men with a range of body sizes. Finally, in this study, stepping responses in forward and backward directions were assessed only with the dominant leg; however, in older adults or people with unilateral impairments (e.g., stroke, Parkinson’s disease), both legs may need to be tested.

This study provides preliminary evidence that the iLEAN test has acceptable inter-rater reliability for assessing reactive balance, with the backward condition demonstrating the highest reliability and sensitivity to change. The iLEAN test may therefore offer physiotherapists a practical, quantitative method for monitoring improvements in reactive balance during exercise or rehabilitation without reliance on costly, laboratory-based equipment. Future research should confirm these findings in older adults and patient populations, and further evaluate intra-rater and test–retest reliability.

## Data Availability

All data produced in the present study are available upon reasonable request to the authors.

## Acknowledgements

We thank Mr Takumi Okumura for their contributions to the data collection. We also thank Mr Yu Imaoka for the development and support of the iLEAN device and application. We thank all participants in the study.

